# Reporting of Retrospective Registration in Clinical Trial Publications: a Cross-Sectional Study of German Trials

**DOI:** 10.1101/2022.10.09.22280784

**Authors:** Martin Haslberger, Stefanie Gestrich, Daniel Strech

## Abstract

**Objective:** Prospective registration has been widely implemented and accepted as a best practice in clinical research, but retrospective registration is still commonly found. We assessed to what extent retrospective registration is reported transparently in journal publications, and investigated factors associated with transparent reporting.

**Design:** We used a dataset of trials registered in ClinicalTrials.gov or Deutsches Register Klinischer Studien, with a German University Medical Center as the lead center, completed 2009–2017, and with a corresponding peer-reviewed results publication. We extracted all registration statements from results publications of retrospectively registered trials and assessed whether they mention or justify the retrospective registration. We analyzed associations of retrospective registration and reporting thereof with registration number reporting, International Committee of Medical Journal Editors (ICMJE) membership/-following and industry sponsorship using chi-squared or Fisher exact test.

**Results:** In the dataset of 1927 trials with a corresponding results publication, 956 (53.7%) were retrospectively registered. Of those, 2.2% (21) explicitly report the retrospective registration in the abstract and 3.5% (33) in the full text. In 2.1% (20) of publications, authors provide an explanation for the retrospective registration in the full text. Registration numbers were significantly underreported in abstracts of retrospectively registered trials compared to prospectively registered trials. Publications in ICMJE member journals did not have statistically significantly higher rates of both prospective registration and disclosure of retrospective registration, and publications in journals claiming to follow ICMJE recommendations showed statistically significantly lower rates compared to non-ICMJE-following journals. Industry sponsorship of trials was significantly associated with higher rates of prospective registration, but not with transparent registration reporting.

**Conclusions:** Contrary to ICMJE guidance, retrospective registration is disclosed and explained only in a small number of retrospectively registered studies. Disclosure of the retrospective nature of the registration would require a brief statement in the manuscript and could be easily implemented by journals.

**Strengths and limitations of this study:**

- We use a large, high-quality dataset of all trials conducted at German university medical centers and registered in two registries, with results publications determined by an extensive manual screening process.
- We consider a period of nine years (2009 – 2017) and describe the development of reporting practices over time
- This study only includes trials led by German university medical centers, reflecting German regulatory standards.

## Introduction

Prospective registration of clinical trials (i.e., registration before enrollment of the first participant) is an important practice to reduce biases in their conduct and reporting (1). A number of ethical and legal documents call for prospective registration: The Declaration of Helsinki (2) and the World Health Organization registry standards (3) state that prospective registration and results reporting of clinical trials are an ethical responsibility. European law, for example, explicitly mandates prospective registration of pharmaceutical trials (4). In addition, many journals, via the International Committee of Medical Journal Editors (ICMJE), encourage or require prospective registration with an appropriate registry before the first participant is enrolled for all trials they publish, as well as the reporting of trial registration numbers (TRNs) in publications for better findability (5,6). Similarly, reporting guidelines such as Consolidated Standards of Reporting Trials (CONSORT) (7) and Good Publication Practice 3 (GPP3) (8) recommend the reporting of trial registration numbers. Prospective registration has been widely implemented and advocated for many reasons: to detect and mitigate publication bias (i.e., the non-reporting of studies, or aspects of studies, that did not yield a positive result) and selective reporting (i.e., the selective reporting of only statistically significant primary outcomes). Prospective registration allows for public scrutiny of trials, identification of research gaps and to support the coordination of efforts by preventing unnecessary duplication (9). When trials are registered retrospectively, i.e., their registry entry is created after study start, this undermines the many of the reasons for registration. While prospective registration has increased over the past decade, retrospective registration is still widespread (10–14). Some registries, such as Deutsches Register Klinischer Studien (DRKS) or the WHO’s International Clinical Trials Registry Platform, explicitly mark retrospectively registered entries as such, whereas others, such as ClinicalTrials.gov, do not. While some journal editors allow retrospectively registered trials to be published, others do not. Journals following ICMJE guidance should in principle mandate prospective registration, but this principle is not always enforced (12,15,16). According to ICMJE guidance, journals should publish retrospectively registered studies only in exceptional cases, noting that “authors should indicate in the publication when registration was completed and why it was delayed. Editors should publish a statement indicating why an exception was allowed.” (5) This was investigated by previous studies which found that such reporting rarely happens (17,18).

Our study aims to investigate the conduct of retrospective registration and its transparent reporting in a larger sample. In a previous study in a cohort of 1509 trials conducted at German University Medical Centers (UMC), registered in DRKS or ClinicalTrials.gov, and reported as complete between 2009-2013, 75% were registered retrospectively (19). This rate dropped to 46% for the 1658 trials completed between 2014-2017 (20). Using the data from these two studies on trials registered in two large registries, led by German UMCs, completed between 2009 and 2017, and with at least one available peer-reviewed results publication (19,20), we investigate whether and how authors report retrospective registration in the results publication. We also explore how retrospective registration is associated with other practices such as TRN reporting.

## Methods

### Data sources and sample

We based our sample on two related projects that were conducted at our research group (19,20). The projects have drawn a full sample (n = 3113) of registry entries for interventional studies reported as complete between 2009 and 2017, led by a German UMC and registered in one of two registries: DRKS, which is the WHO primary trial registry for Germany, and ClinicalTrials.gov, which is also routinely used in Germany to register clinical research and accepted by the ICMJE. Our dataset also includes the earliest results publications found for 68.4% (2129/3113) of the trials, which were manually identified in different stages until September 1st, 2020. We retrieved the combined data from the two projects from a GitHub repository (https://github.com/maia-sh/intovalue-data, accessed 22.02.2022). The final dataset is publicly available (21).

### Eligibility criteria

We included any trial that [1] was registered as an interventional study in either the ClinicalTrials.gov or the DRKS database, [2] was completed between 2009 and 2017, [3] reports a German UMC listed as the responsible party or lead sponsor, or with a principal investigator from a German UMC, [4] has published results in a peer-reviewed journal. Detailed descriptions of how these variables were derived are provided in the original publications of the dataset (19,20). Retrospective registration was determined based on the registration and study start dates in the registry entries: dates were set to the first of the respective month and studies with a registration date more than one month after start date counted as retrospectively registered. For trials that were registered in both registries, we kept the entry that was created earlier.

### Data extraction

For all retrospectively registered trials, we manually searched the abstract and the full text of the publications, including editorial statements, whether they reported

- the fact that the study was registered (binary),
- a trial registration number (binary),
- the exact wording used to report the registration, including any provided registration numbers (free text),
- the date of the retrospective registration (binary), and
- the fact that the study was retrospectively registered (binary).
- We also assessed whether (binary) and how authors justified or explained the retrospective registration (free text).

One rater (MH) used the keywords “regist”, “nct”, “drks”, “eudra”, “retro”, “delay”, and “after” to search for registration numbers and wording pointing to retrospective registration in all publications. We considered a retrospective registration statement transparent if the authors explicitly mentioned that the registration was retrospective, e.g., “this study was retrospectively registered in [registry], [TRN]”. Reporting of the registration date alone was not considered as transparent reporting of retrospective registration, except if the date of registration was mentioned in combination with the study start date in the same paragraph.

### ICMJE journals

We created additional variables for whether journals are ICMJE members or follow the ICMJE recommendations (22).

### Cross-registrations

We classified all retrospectively registered studies in our sample that also report a registration in EudraCT in the publication as prospective, as registrations on the platform are required prior to the approval of regulatory agencies or research ethics committees (4).

### Reliability assessment of ratings

To assess the reliability of the data extraction, another rater (SG) performed three validation steps: first, a sample of 100 publications was screened using the same extraction form, during the main screening to refine category definitions. Second, another sample of 100 publications for which no registration number reporting was noted by MH to check for false negative ratings. Third, all cases with either date, or reporting of retrospective registration or justification were screened, to check for false positives.

### Analyses

#### Associations between prospective registration and other variables

To test the strength of the associations between prospective registration and three variables, we used Pearson’s chi-squared independence test. These variables were (1) publication in a ICMJE member journal or a journal following ICMJE recommendations, (2) reporting of a registration number, and (3) industry funding.

#### Associations between reporting of retrospective registration and other variables

To test the strength of the associations between the reporting of retrospective registration and two binary variables, we used Fisher’s exact test, as case numbers were low. These variables are (1) publication in a ICMJE member journal or a journal following ICMJE recommendations, and (2) industry funding.

#### Software

We used Microsoft Excel for data collection and R (version 4.0.3) for data analysis and visualization.

#### Reporting

We checked our manuscript against the STROBE checklist (supplementary Table 1) (23).

#### Patient and Public Involvement

No patient involved.

## Results

### Sample of retrospectively registered trials

After applying the above-mentioned exclusion criteria, 1932 registered studies with an associated results publication remained. Of these, 1038 (54%) were retrospectively registered according to the information provided in ClinicalTrials.gov and DRKS. We screened these 1038 studies for our analysis. Five of the publications were excluded as they were mislabeled as results publications in the dataset. Another 77 (8%) of the publications provided a EudraCT number, in which case we reclassified the study as prospectively registered, leaving 956 studies. For statistical comparisons, we used the studies classified as prospectively registered (n=971) in the dataset as a control group. A flowchart of this study selection is provided in Figure 1. Basic characteristics of included trials are available in supplementary Table 2.

**Figure 1:**
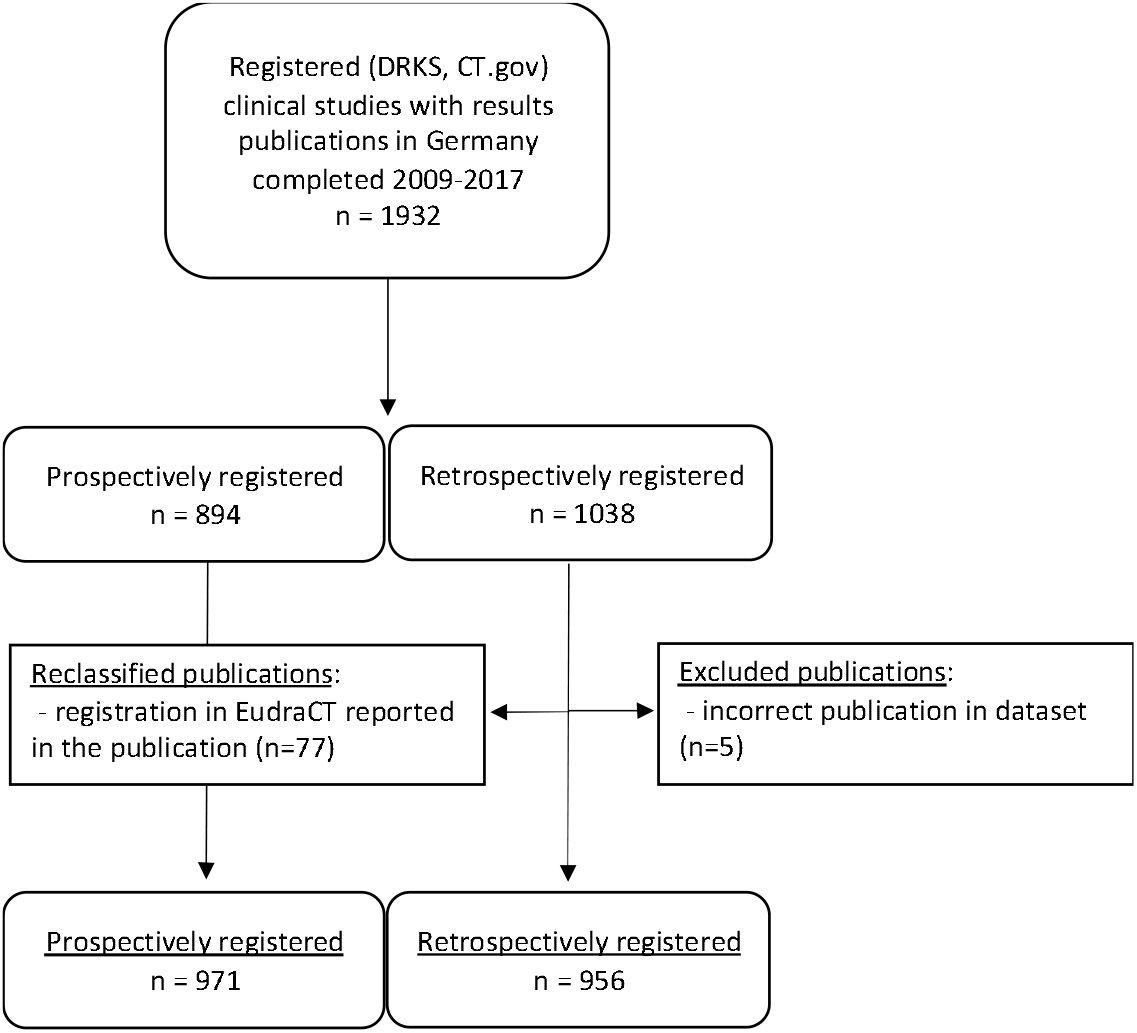
Flowchart of in-/exclusion of studies. From the 1038 trials that were retrospectively registered in Clincialtrials.gov or DRKS, we excluded 5 publications that clearly did not report clinical study results (e.g., secondary analyses of CT data) and another 77 that reported EudraCT entries in the publications, resulting in 956 retrospectively registered studies from a total dataset of 1927 (971 + 956) studies.

### Retrospective registration

Figure 2 shows the extent of retrospective registration over time, which has been falling steadily from 100% in 2004 to 25% in 2017.

**Figure 2:**
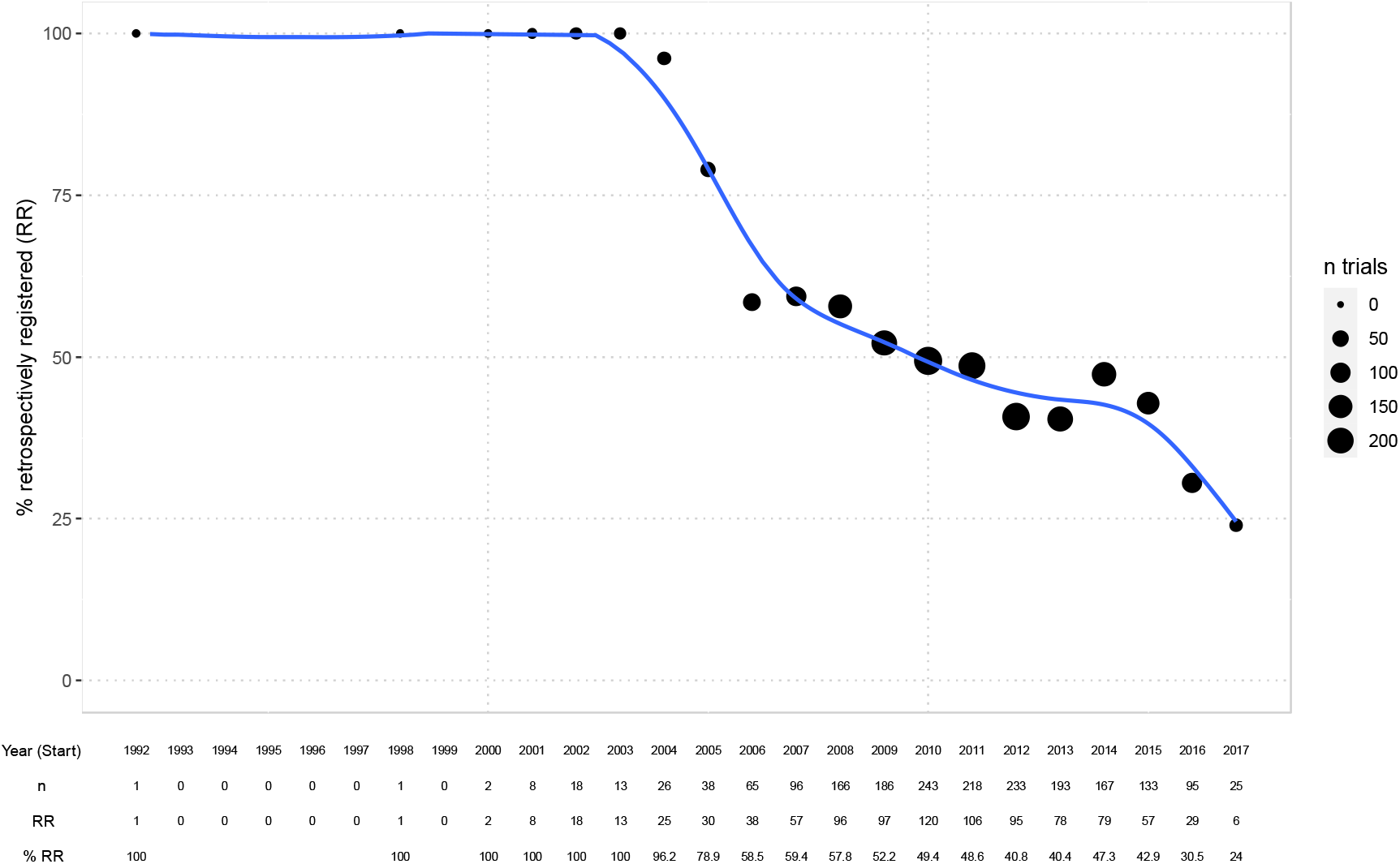
Percentage of retrospectively registered (RR) trials over time (per study start year). GAM (generalized additive model) smoother laid over (blue) with 95% confidence interval. Bubble sizes indicate the number of trials per year included in the dataset.

We describe associations between prospective registration and previously defined binary variables in Table 1 We found no statistically significant association between publication in ICMJE member journals and prospective registration (p=0.10). Similarly, we found no statistically significant association with prospective registration when also including publication in journals reporting to follow ICMJE recommendations (p=0.47). It is important to note here that the information on ICMJE-following is based on journals’ requests to be included on the ICMJE website as a journal following the ICMJE’s recommendations (22), therefore our results suggest that journals requesting to be listed on the site often do not enforce the recommendations strongly. However, there are other journals, such as many PLOS journals, that are not featured on the ICMJE site, but implement the recommendations. Retrospectively registered trials, compared to prospectively registered trials significantly underreported registration numbers in the abstract (p = 0.0007). Industry sponsorship of trials was associated with prospective registration (p = 0.002). In 31% (294/956) of trials, registration occurred between study completion and publication (median 370 days before publication). Another 3% (25/956) of trials were registered after publication (median 249 days after publication).

**Table 1:**
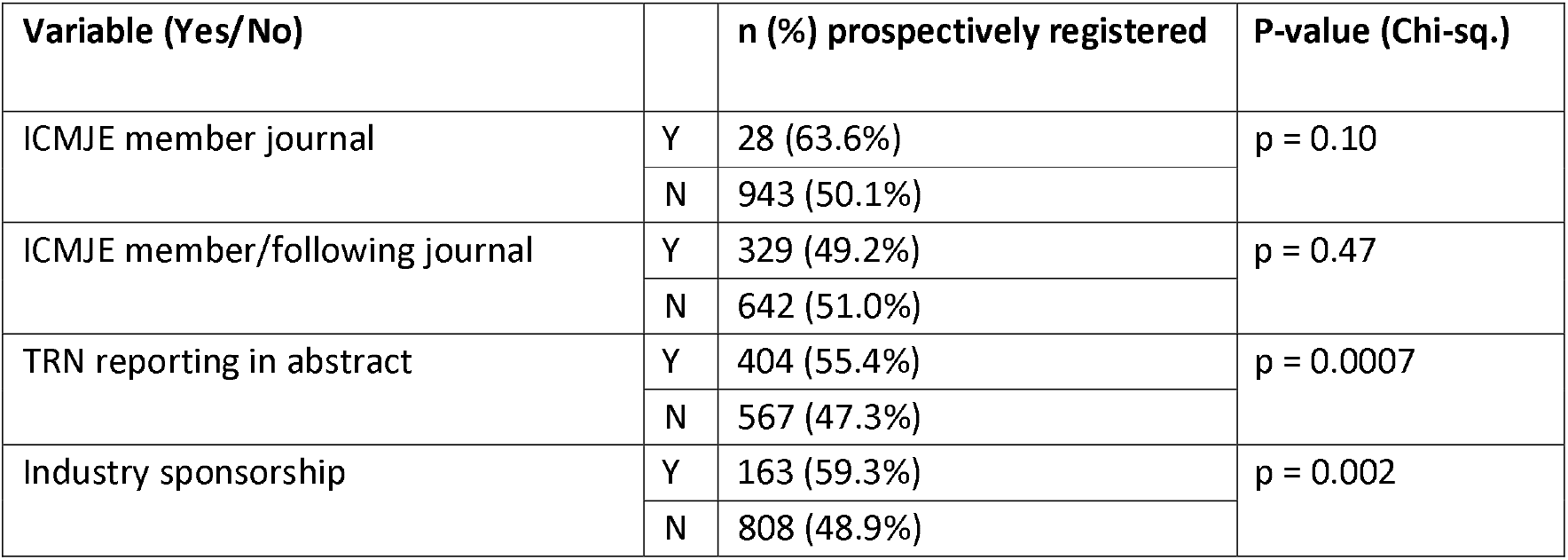
Associations between prospective registration and other variables

| Variable (Yes/No) |  | n (%) prospectively registered | P-value (Chi-sq.) |
| --- | --- | --- | --- |
| ICMJE member journal | Y | 28 (63.6%) | p = 0.10 |
|  | N | 943 (50.1%) |  |
| ICMJE member/following journal | Y | 329 (49.2%) | p = 0.47 |
|  | N | 642 (51.0%) |  |
| TRN reporting in abstract | Y | 404 (55.4%) | p = 0.0007 |
|  | N | 567 (47.3%) |  |
| Industry sponsorship | Y | 163 (59.3%) | p = 0.002 |
|  | N | 808 (48.9%) |  |

### Reporting of registration

Table 2 summarizes the prevalence of reporting of trial registration and the reporting of retrospective registration. In 82% (783/956) of the remaining results publications of retrospectively registered trials, the registration was explicitly reported in either the abstract or the full text. In all except four of these publications, the registration was mentioned by providing the registration number. In the other cases, the registration was mentioned but without reporting a registration number.

**Table 2:** Number of retrospectively registered trials and prevalence of key retrospective registration reporting practices. * “other” includes footnotes, sidebars, etc.

|  | n | % (of total) |
| --- | --- | --- |
| **Total: Retrospectively registered trials** | **956** | **100.0%** |
| Registration reported | 783 | 81.9% |
| **Registration number reported** | **779** | **81.5%** |
| *in abstract* | 325 | 34.0% |
| *in full-text* | 535 | 56.0% |
| *in other\** | 134 | 14.0% |
| **Registration date reported** | **67** | **7.0%** |
| *in abstract* | 45 | 4.7% |
| *in full-text* | 32 | 3.3% |
| **Retrospective registration addressed** | **47** | **4.9%** |
| *in abstract* | 21 | 2.2% |
| *in full-text* | 33 | 3.5% |
| **Retrospective registration justified/explained** | **20** | **2.1%** |
| *in abstract* | 0 | 0.0% |
| *in full-text* | 20 | 2.1% |

### Reporting of retrospective registration

The rate of trials for which retrospective registration is reported transparently increased over the last years up to 15% in 2020 (Figure 3). Overall, among all 956 retrospectively registered clinical studies, five percent (47) mention explicitly that this registration was retrospective in the abstract or full text (see Table 2). Among those cases, 20 give some explanation or justification for why registration was retrospective. In seven percent (67) of cases, the authors reported the registration date alongside the registration statement, but in 35 of those, the date was provided without giving the necessary context that the registration was retrospective.

**Figure 3:**
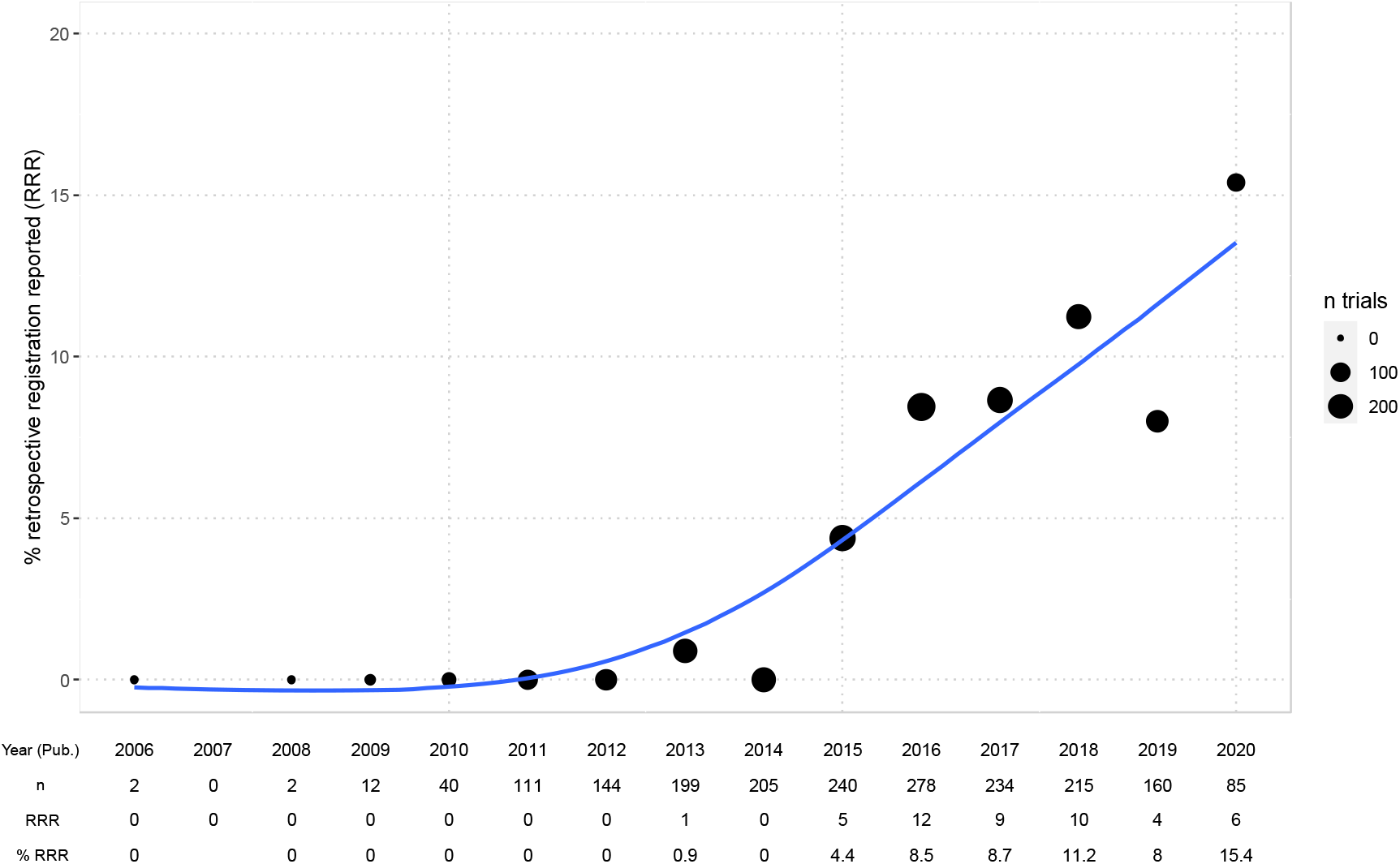
Percentage of retrospectively registered trials reporting retrospective registration transparently in the publication over time (per study publication year). GAM (generalized additive model) smoother laid over (blue) with 95% confidence interval. Bubble sizes indicate the number of trials per year included in the dataset. Starting in 2013, some authors begin to report retrospective registration. 15% of publications of retrospectively registered trials from 2020 transparently report retrospective registration. Four trials were published before 2009 – in all those cases the study completion dates provided in the registry were after 2009. Study start dates were before 2005 and studies were registered in 2005 (3/4) or later (1/4).

Publications in ICMJE member journals did not have a statistically significantly higher rate of reporting of retrospective registration (13% vs. 5%, p = 0.18), whereas publications in ICMJE member or -following journals had a significantly lower rate (2% vs. 7%, p = 0.004). We found no association with transparent reporting of retrospective registration for industry sponsored trials (2% vs. 5%, p = 0.16) (Table 3).

**Table 3:**
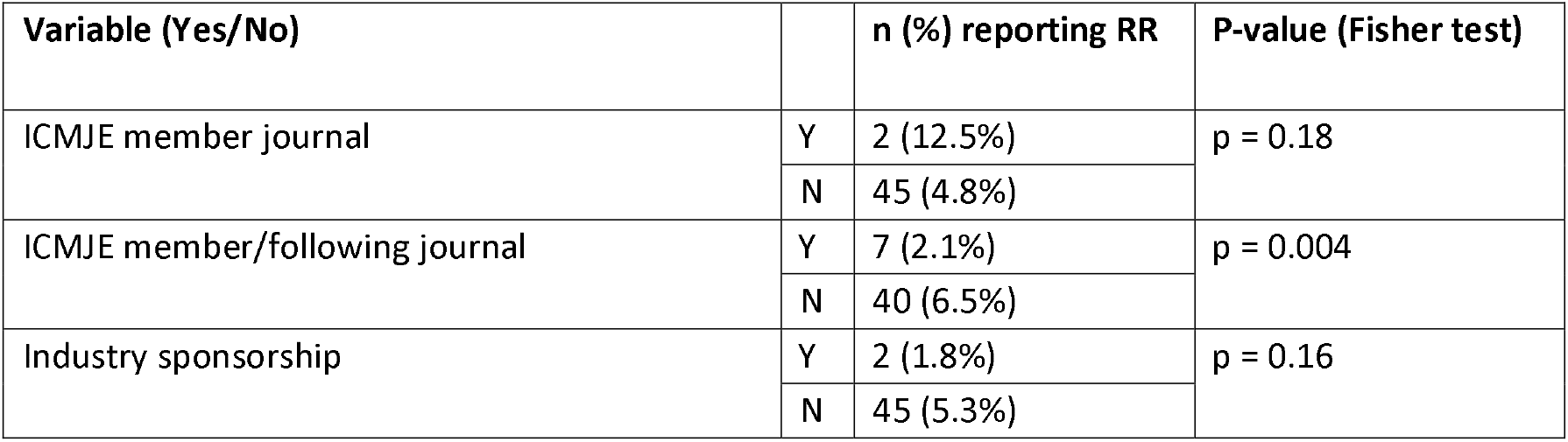
Associations between transparent reporting of retrospective registration and other variables

| Variable (Yes/No) |  | n (%) reporting RR | P-value (Fisher test) |
| --- | --- | --- | --- |
| ICMJE member journal | Y | 2 (12.5%) | $p = 0.18$ |
|  | N | 45 (4.8%) |  |
| ICMJE member/following journal | Y | 7 (2.1%) | $p = 0.004$ |
|  | N | 40 (6.5%) |  |
| Industry sponsorship | Y | 2 (1.8%) | $p = 0.16$ |
|  | N | 45 (5.3%) |  |

### Justifications of retrospective registration

In 20 cases in which the retrospective nature of the registration was reported, the authors provided further information explaining or justifying the retrospective registration. Notably, 14 of the 20 studies (70%) that justified the retrospective registration were published in a single journal, PLOS ONE. Table 4 shows the main themes present in authors’ explanations, with text examples.

**Table 4.**
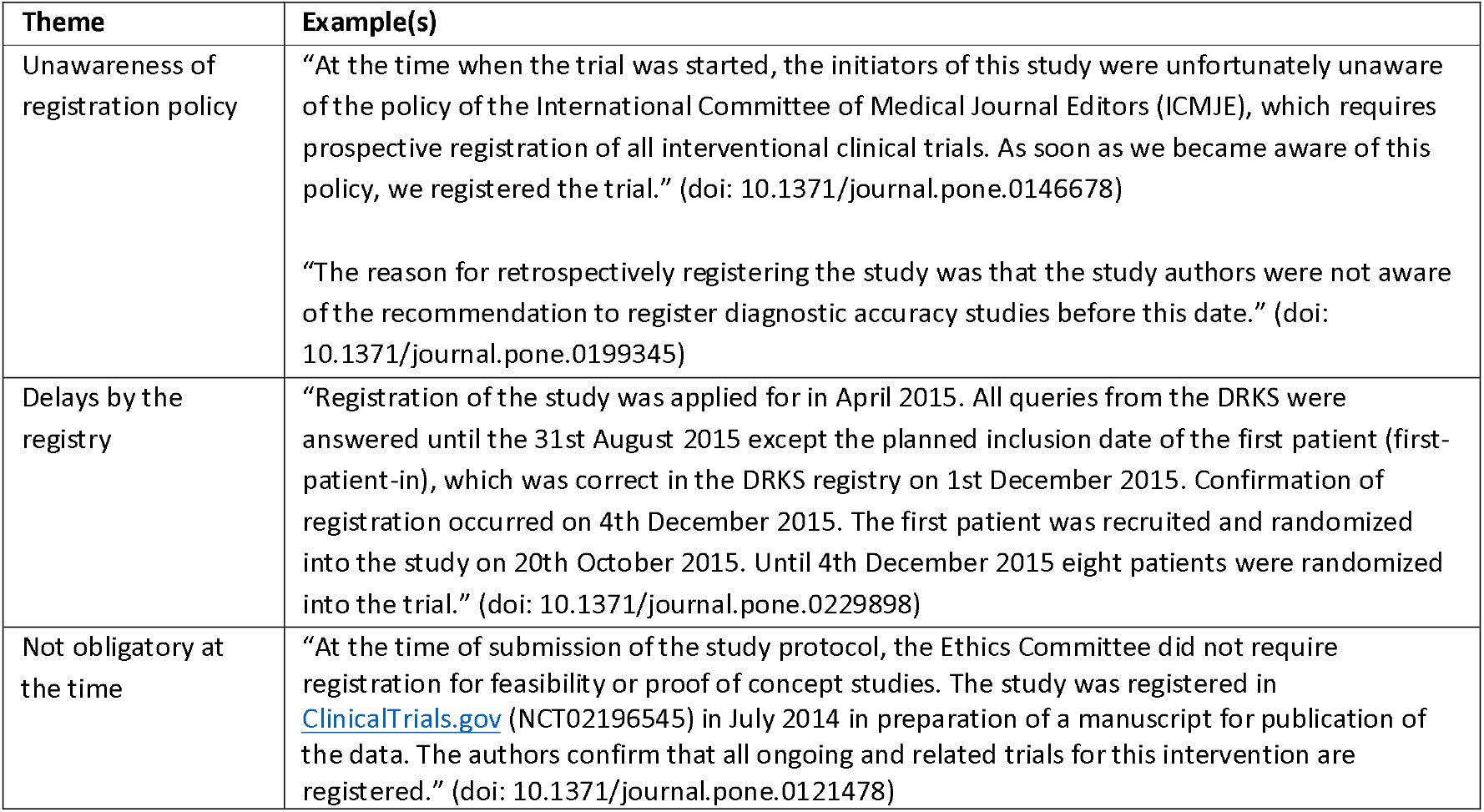
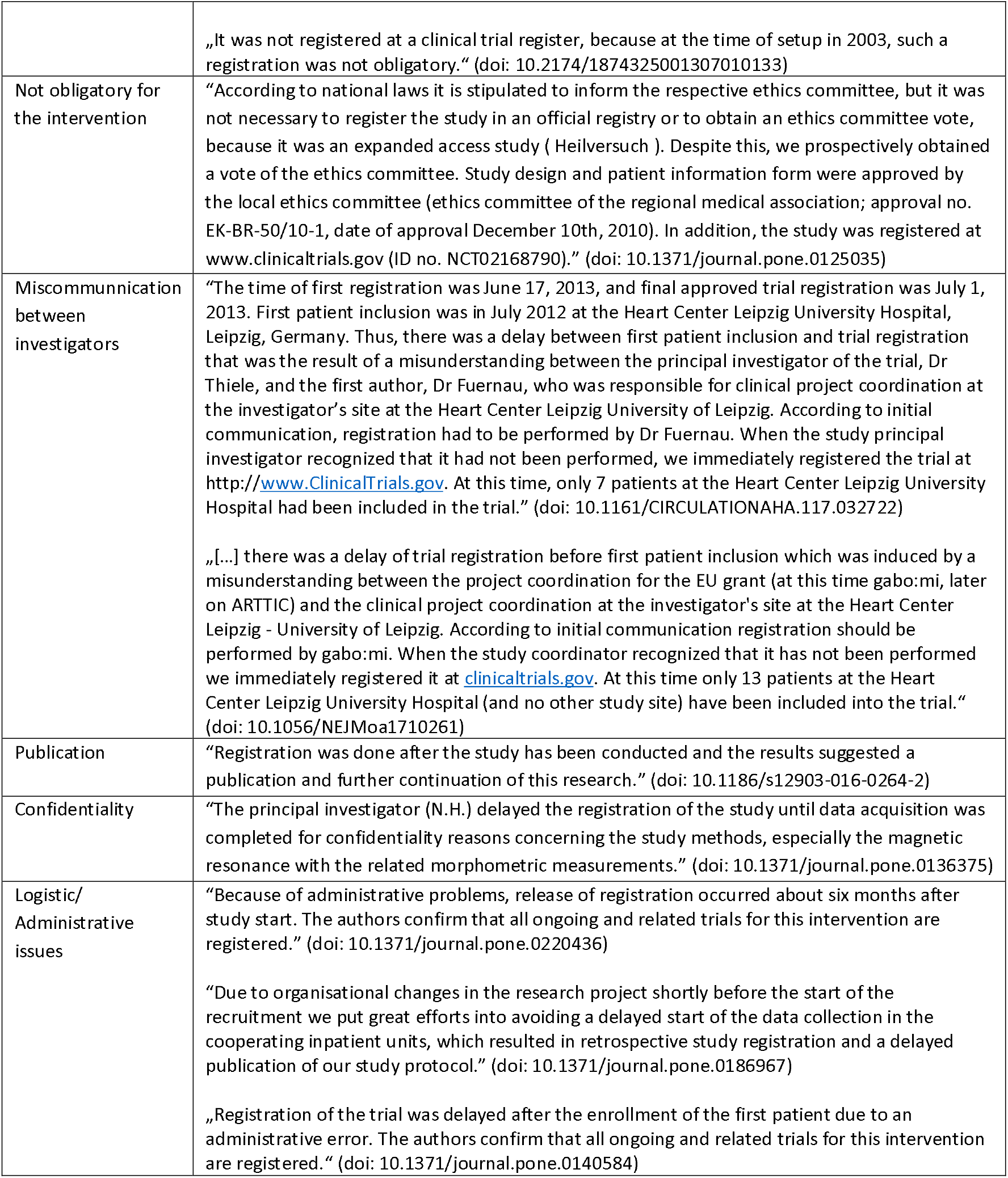
Main themes identified from authors’ explanations of retrospective reporting and example statements.

## Discussion

In this study we show that in a sample of 956 results publications from retrospectively registered clinical studies led by German UMCs and completed between 2009 and 2017, only a small number of publications (5%) make the retrospective nature of the registration transparent, and even fewer (2%) explain the reasons for retrospective registration. To our knowledge, two studies have previously quantified the reporting of retrospective registration: Al-Durra et al. (17) found in a sample of 286 publications in ICMJE member journals and published in 2018 that only three percent (8/286) of papers of retrospectively registered trials in their sample include justifications or explanations for delayed registration. Similarly, Loder et al. (18), in their analysis of 70 papers submitted to the British Medical Journal from 2012-2015 and rejected for registration issues, found that three percent (2/70) disclosed the registration problem when published in another journal. Our study finds a slightly lower percentage of two percent for explanations of the reasons for retrospective registration, but a higher percentage of five percent for disclosure in a larger sample representing a broader selection of journals and extended time frame.

We found that publications were not significantly more often prospectively registered when they were published in ICMJE member journals or in journals following ICMJE recommendations, but showed a significantly higher rate of TRN reporting. A similar result was found by Al-Durra et al. (17). Further, we found that transparent reporting of retrospective registration does not happen significantly more often in publications in ICMJE member journals, and is even happening at a significantly lower rate in journals listed as following ICMJE recommendations.

There were different reasons for retrospective registration brought forth by authors, many of which have been described previously (15,17,18,24). In some cases, authors raise points that lie outside their direct responsibility, such as delays caused by the registry or research not being legally required to be preregistered. Several other reasons provided were within authors’ control, such as logistic and administrative issues, miscommunication between researchers or unawareness of registration policies. In some cases, authors report registering a study to meet journal editorial policies even though registration would not be required for the kind of research otherwise. This is also possibly reflected in the fact that almost a third (31%) of retrospectively registered studies in our sample have been registered between study completion and publication. In one publication, the authors transparently describe that the registration occurred only when “results suggested a publication and further continuation of this research”, which has been previously described as “selective registration bias” (17) and is explicitly called out in ICMJE guidance as it “meets none of the purposes of preregistration” (5). Another identified theme revolves around the confidentiality of methods – however, in this case many other details about the trial could have been preregistered.

### Limitations

For feasibility and data quality reasons, our study was based on an existing validated dataset, containing only trials led by German UMCs, which might limit its generalizability to other regions. However, the sample also contained multi-center trials with other countries involved and is larger and from a wider variety of journals compared to previous studies (17,18). Our analysis of retrospective registration is based on trial start dates and registration dates as provided by the two registries used for sampling: Clinicaltrials.gov and DRKS. It is possible that authors did not update their registry entries when delays to the start date occurred. For example, we did not specifically follow up cases in which authors wrote that a trial was registered prospectively, but the registry dates did not reflect that statement. In order not to reduce the sample size, we also did not correct for varying follow-up in the identification of result publications, e.g., by limiting our analysis to publications published within 2 years of trial completion. However, this means that the newer trials in the sample (i.e., years 2016, 2017) might not reflect the complete research output of those years as some trials may not have been published by the end of follow-up in 2020 and were therefore excluded from the analysis. The numbers presented in Figures 2 and 3 may overestimate the improvements in prospective registration as trials reporting results on time might likely generally show a higher quality of registration conduct and might therefore be registered prospectively at a higher rate.

In our analyses involving the classification into ICMJE-following and non-following journals, we relied on the data provided on the ICMJE website (icmje.org), which are self-reported by journals, i.e., a journal must write to the ICMJE that they want to be included in the list. Thus, there are some journals missing in the ICMJE data and therefore in our dataset. For ICMJE member journals (n=12) on the other hand, there is a complete listing available.

### Conclusion

The Declaration of Helsinki and other guidelines for responsible clinical research unanimously recommend prospective registration of all clinical research (2). For clinical trials regulated by drug and device regulatory authorities, this was codified into law (4). A major aim of prospective registration is to minimize the risk of undisclosed changes to the protocol after the study started and first results are analyzed. When registration happens retrospectively, this major goal is not addressed. The reporting of study registration is generally considered a best practice to make a study more trustworthy. In the case of retrospective registration, in contrast, reporting registration without transparency on the retrospective nature should rather raise concerns as readers might wrongly interpret the mentioning of registration as a quality criterion. This could be considered “performative reproducibility”, i.e., the “pretence of reproducibility without the reality” (25). Journal editors and reviewers could enforce explicit reporting and explanation of retrospective registration, but we found that this rarely happens. To fulfill the ICMJE requirements on reporting retrospective registration, a simple note in the registration statement of the paper would suffice, such as: “This study was retrospectively registered as [TRN] at [Registry], [X] days after the trial started because [Reason]”.

## Data Availability

All code and the data for this study are available at https://github.com/mhaslberger/retrospective-registration.
Data are also available in an OSF repository (https://osf.io/8g5cf/).

## Ethics approval

Not applicable

## Contributorship statement

Martin Haslberger – Conceptualization, methodology, investigation, analysis, writing - original draft, project management

Stefanie Gestrich – Methodology, investigation

Daniel Strech – Conceptualization, methodology, supervision, writing – review and editing, funding acquisition

## Data sharing statement

All code and the data for this study are available at https://github.com/mhaslberger/retrospective-registration. Data are also available in an OSF repository (https://osf.io/8g5cf/).

## Acknowledgments

We thank Maia Salholz-Hillel for conceptual feedback and technical support. We thank Martin R. Holst and Dr. Delwen Franzen for feedback on the manuscript.

## References

1. Simes RJ. Publication bias: the case for an international registry of clinical trials. J Clin Oncol. 1986 Oct;4(10):1529–41.

2. WMA. World Medical Association: Declaration of Helsinki - Ethical Principles for Medical Research involving Human Subjects [Internet]. 2018. Available from: https://www.wma.net/policies-post/wma-declaration-of-helsinki-ethical-principles-for-medical-research-involving-human-subjects/

3. World Health Organization. International standards for clinical trial registries: the registration of all interventional trials is a scientific, ethical and moral responsibility [Internet]. version 3.0. Geneva: World Health Organization; 2018 [cited 2022 Feb 1]. 48 p. Available from: https://apps.who.int/iris/handle/10665/274994

4. European Medicines Agency. Regulation (EU) No 536/2014 of the European Parliament and of the Council of 16 April 2014 on clinical trials on medicinal products for human use, and repealing Directive 2001/20/EC [Internet]. Jan 31, 2022. Available from: http://data.europa.eu/eli/reg/2014/536/2022-01-31

5. ICMJE. International Committee of Medical Journal Editors Recommendations for the Conduct, Reporting, Editing, and Publication of Scholarly Work in Medical Journals [Internet]. [cited 2021 Oct 18]. Available from: http://www.icmje.org/icmje-recommendations.pdf

6. De Angelis C, Drazen JM, Frizelle FA, Haug C, Hoey J, Horton R, et al. Clinical trial registration: a statement from the International Committee of Medical Journal Editors. Lancet Lond Engl. 2004 Sep 11;364(9438):911–2.

7. Moher D, Hopewell S, Schulz KF, Montori V, Gotzsche PC, Devereaux PJ, et al. CONSORT 2010 Explanation and Elaboration: updated guidelines for reporting parallel group randomised trials. BMJ. 2010 Mar 23;340(mar23 1):c869–c869.

8. Battisti WP, Wager E, Baltzer L, Bridges D, Cairns A, Carswell CI, et al. Good Publication Practice for Communicating Company-Sponsored Medical Research: GPP3. Ann Intern Med. 2015 Sep 15;163(6):461–4.

9. Zarin DA, Keselman A. Registering a Clinical Trial in ClinicalTrials.gov. Chest. 2007 Mar;131(3):909–12.

10. Trinquart L, Dunn AG, Bourgeois FT. Registration of published randomized trials: a systematic review and meta-analysis. BMC Med. 2018 Oct 16;16(1):173.

11. Birajdar AR, Bose D, Nishandar TB, Shende AA, Thatte UM, Gogtay NJ. An audit of studies registered retrospectively with the Clinical Trials Registry of India: A one year analysis. Perspect Clin Res. 2019 Mar;10(1):26–30.

12. Scott A, Rucklidge JJ, Mulder RT. Is Mandatory Prospective Trial Registration Working to Prevent Publication of Unregistered Trials and Selective Outcome Reporting? An Observational Study of Five Psychiatry Journals That Mandate Prospective Clinical Trial Registration. Wicherts JM, editor. PLOS ONE. 2015 Aug 19;10(8):e0133718.

13. Mann E, Nguyen N, Fleischer S, Meyer G. Compliance with trial registration in five core journals of clinical geriatrics: a survey of original publications on randomised controlled trials from 2008 to 2012. Age Ageing. 2014 Nov;43(6):872–6.

14. Smaïl-Faugeron V, Fron-Chabouis H, Durieux P. Clinical trial registration in oral health journals. J Dent Res. 2015 Mar;94(3 Suppl):8S–13S.

15. Harriman SL, Patel J. When are clinical trials registered? An analysis of prospective versus retrospective registration. Trials. 2016 Apr 15;17:187.

16. Boughton S. Retrospectively registered trials: the Editors’ dilemma [Internet]. Research in progress blog. 2016 [cited 2022 Jul 21]. Available from: https://blogs.biomedcentral.com/bmcblog/2016/04/15/retrospectively-registered-trials-editors-dilemma/

17. Al-Durra M, Nolan RP, Seto E, Cafazzo JA. Prospective registration and reporting of trial number in randomised clinical trials: global cross sectional study of the adoption of ICMJE and Declaration of Helsinki recommendations. BMJ. 2020 Apr 14;m982.

18. Loder E, Loder S, Cook S. Characteristics and publication fate of unregistered and retrospectively registered clinical trials submitted to The BMJ over 4 years. BMJ Open. 2018 Feb 16;8(2):e020037.

19. Wieschowski S, Riedel N, Wollmann K, Kahrass H, Müller-Ohlraun S, Schürmann C, et al. Result dissemination from clinical trials conducted at German university medical centers was delayed and incomplete. J Clin Epidemiol. 2019 Nov;115:37–45.

20. Riedel N, Wieschowski S, Bruckner T, Holst MR, Kahrass H, Nury E, et al. Results dissemination from completed clinical trials conducted at German university medical centers remained delayed and incomplete. The 2014 –2017 cohort. J Clin Epidemiol. 2022 Apr;144:1–7.

21. [dataset] Haslberger M, Gestrich S, Strech D. Reporting of Retrospective Registration in Clinical Trial Publications [Internet]. 2023. Available from: https://osf.io/8g5cf/

22. ICMJE. Journals stating that they follow the ICMJE Recommendations [Internet]. [cited 2022 Apr 7]. Available from: http://www.icmje.org/journals-following-the-icmje-recommendations/

23. von Elm E, Altman DG, Egger M, Pocock SJ, Gøtzsche PC, Vandenbroucke JP, et al. The Strengthening the Reporting of Observational Studies in Epidemiology (STROBE) statement: guidelines for reporting observational studies. Lancet Lond Engl. 2007 Oct 20;370(9596):1453–7.

24. Hunter KE, Seidler AL, Askie LM. Prospective registration trends, reasons for retrospective registration and mechanisms to increase prospective registration compliance: descriptive analysis and survey. BMJ Open. 2018 Mar 1;8(3):e019983.

25. Buck S. Beware performative reproducibility. Nature. 2021 Jul 8;595(7866):151–151.

