## supplementary Table 1 for "Reporting of Retrospective Registration in Clinical Trial Publications: a Cross-Sectional Study of German Trials"

Supplementary Table 1: STROBE checklist

| | Item No. | Recommendation | Page No. | Relevant text from manuscript |
|---|---|---|---|---|
| **Title and abstract** | 1 | (*a*) Indicate the study's design with a commonly used term in the title or the abstract | 2 | Cross-sectional study |
| | | (*b*) Provide in the abstract an informative and balanced summary of what was done and what was found | 2 | See abstract |
| **Introduction** | | | | |
| Background/rationale | 2 | Explain the scientific background and rationale for the investigation being reported | 3 | When trials are registered retrospectively, i.e., their registry entry is created after study start, this undermines the many of the reasons for registration. While prospective registration has increased over the past decade, retrospective registration is still widespread (10–14). Some registries, such as DRKS, explicitly mark retrospectively registered entries as such, whereas others, such as ClinicalTrials.gov, do not. While some publishers allow retrospectively registered trials to be published, others do not. Journals following ICMJE guidance should in principle mandate prospective registration, but this principle is not always enforced (12,15,16). |
| Objectives | 3 | State specific objectives, including any prespecified hypotheses | 3 | Our study aims to investigate the conduct of retrospective registration and its transparent reporting |
| **Methods** | | | | |
| Study design | 4 | Present key elements of study design early in the paper | 3 | We based our sample on two related projects that were conducted at our research group (17,18). The projects have drawn a full sample (n = 3113) of registry entries for interventional studies reported as complete between 2009 and 2017, led by a German University Medical Center and registered in one of two registries: The ClinicalTrials.gov platform (CT.gov) and the Deutsches Register Klinischer Studien (DRKS) |
| Setting | 5 | Describe the setting, locations, and relevant dates, including periods of recruitment, exposure, follow-up, and data collection | 3-4 | See above |

| | | | | |
|---|---|---|---|---|
| Participants | 6 | *Cross-sectional study*—Give the eligibility criteria, and the sources and methods of selection of participants | 3-4 | We included any trial that [1] was registered as an interventional study in either the ClinicalTrials.gov or the DRKS database, [2] was completed between 2009 and 2017, [3] reports a German University Medical Center (UMC) listed as the responsible party or lead sponsor, or with a principal investigator from a German UMC, [4] has published results in a peer-reviewed journal. |
| Variables | 7 | Clearly define all outcomes, exposures, predictors, potential confounders, and effect modifiers. Give diagnostic criteria, if applicable | 3 | we investigate whether and how authors report retrospective registration in the results publication. We also explore trends over time and how retrospective registration is associated with other practices such as reporting the trial registration number. |
| Data sources/ measurement | 8* | For each variable of interest, give sources of data and details of methods of assessment (measurement). Describe comparability of assessment methods if there is more than one group | 3-4 | Data sources and sample. We based our sample on two related projects that were conducted at our research group (17,18). The projects have drawn a full sample (n = 3113) of registry entries for interventional studies reported as complete between 2009 and 2017, led by a German University Medical Center and registered in one of two registries: The ClinicalTrials.gov platform (CT.gov) and the Deutsches Register Klinischer Studien (DRKS), which is the WHO primary trial registry for Germany. Our dataset also includes the earliest results publications found for 68.4% (2129/3113) of the trials, which were manually identified in different stages until September 1st, 2020. We retrieved the combined data from the two projects from a GitHub repository (https://github.com/maia-sh/intovalue-data, accessed 22.02.2022). |
| Bias | 9 | Describe any efforts to address potential sources of bias | 4 | *Reliability assessment of ratings.* To assess the reliability of the data extraction, another rater (SG) performed three validation steps: first, a sample of 100 publications was screened using the same extraction form, during the main screening to refine category definitions. Second, another sample of 100 publications for which no registration number reporting was noted by MH to check for false negative ratings. Third, all cases with either date, or reporting of retrospective registration or justification were screened, to check for false positives. |

| | | | | |
|---|---|---|---|---|
| Study size | 10 | Explain how the study size was arrived at | 3 | We based our sample on two related projects that were conducted at our research group |
| Quantitative variables | 11 | Explain how quantitative variables were handled in the analyses. If applicable, describe which groupings were chosen and why | - | Not applicable |
| Statistical methods | 12 | (*a*) Describe all statistical methods, including those used to control for confounding | 4 | See Methods |
| | | (*b*) Describe any methods used to examine subgroups and interactions | 4 | See Methods |
| | | (*c*) Explain how missing data were addressed | - | Not applicable |
| | | (*d*) *Cross-sectional study*—If applicable, describe analytical methods taking account of sampling strategy | - | Not applicable |
| | | (*e*) Describe any sensitivity analyses | - | Not applicable |
| **Results** | | | | |
| Participants | 13* | (a) Report numbers of individuals at each stage of study—eg numbers potentially eligible, examined for eligibility, confirmed eligible, included in the study, completing follow-up, and analysed | 5 | First paragraph of Results |
| | | (b) Give reasons for non-participation at each stage | 5 | First paragraph of Results |
| | | (c) Consider use of a flow diagram | 5 | Figure 1 |
| Descriptive data | 14* | (a) Give characteristics of study participants (eg demographic, clinical, social) and information on exposures and potential confounders | 5 | Supplementary table 1 |
| | | (b) Indicate number of participants with missing data for each variable of interest | - | Complete case analysis |
| | | (c) *Cohort study*—Summarise follow-up time (eg, average and total amount) | 3 | The projects have drawn a full sample (n = 3113) of registry entries for interventional studies reported as complete between 2009 and 2017, led by a German University Medical Center and registered in one of two registries: The ClinicalTrials.gov platform (CT.gov) and the Deutsches Register Klinischer Studien (DRKS), which is the WHO primary trial registry for Germany. Our dataset also includes the earliest results publications found for 68.4% (2129/3113) of the trials, which were manually identified in different stages until September 1st, 2020 |
| Outcome data | 15* | *Cross-sectional study*—Report numbers of outcome events or summary measures | *5-7* | *See results, e.g. Table 2, Figures 2,3* |
| Main results | 16 | (*a*) Give unadjusted estimates and, if applicable, confounder-adjusted estimates and | - | Not applicable |

| | | | | |
|---|---|---|---|---|
| | | their precision (eg, 95% confidence interval). Make clear which confounders were adjusted for and why they were included | | |
| | | (*b*) Report category boundaries when continuous variables were categorized | - | Not applicable |
| | | (*c*) If relevant, consider translating estimates of relative risk into absolute risk for a meaningful time period | - | Not applicable |
| Other analyses | 17 | Report other analyses done—eg analyses of subgroups and interactions, and sensitivity analyses | 6-9 | See results |
| **Discussion** | | | | |
| Key results | 18 | Summarise key results with reference to study objectives | 9-10 | See 1st paragraph of discussion |
| Limitations | 19 | Discuss limitations of the study, taking into account sources of potential bias or imprecision. Discuss both direction and magnitude of any potential bias | 10 | See Limitations section |
| Interpretation | 20 | Give a cautious overall interpretation of results considering objectives, limitations, multiplicity of analyses, results from similar studies, and other relevant evidence | 9-10 | See first paragraph of discussion |
| Generalisability | 21 | Discuss the generalisability (external validity) of the study results | 10 | For feasibility and data quality reasons, our study was based on an existing validated dataset, containing only trials led by German UMCs, which might limit its generalizability to other regions. […] |
| **Other information** | | | | |
| Funding | 22 | Give the source of funding and the role of the funders for the present study and, if applicable, for the original study on which the present article is based | 1 | This work was partly funded under a grant from the Federal Ministry of Education and Research of Germany (Bundesministerium fuer Bildung und Forschung - BMBF) [01PW18012]. The funder was not involved in the study design, data collection, analysis, or interpretation, writing of the manuscript, or the decision to submit for publication. |
