## supplementary Table 2 for "Reporting of Retrospective Registration in Clinical Trial Publications: a Cross-Sectional Study of German Trials"

Supplementary Table 2: Basic characteristics of included trials

| | **Retrospectively registered trials (n = 956)** | **Prospectively registered trials (n = 971)** |
|---|---|---|
| **Registry** | | |
| ClinicalTrials.gov | 713 | 766 |
| DRKS | 243 | 205 |
| **Sponsorship** | | |
| Industry | 112 | 163 |
| Other | 844 | 808 |
| **Phase** | | |
| Phase 1 | 33 | 71 |
| Phase 2 | 122 | 206 |
| Phase 3 | 91 | 134 |
| Phase 4 | 78 | 99 |
| No phase | 632 | 461 |
| **Intervention type** | | |
| Behavioral | 94 | 72 |
| Biological | 18 | 32 |
| Device | 199 | 138 |
| Dietary Supplement | 40 | 30 |
| Drug | 186 | 340 |
| Genetic | 1 | 1 |
| Procedure | 96 | 70 |
| Radiation | 6 | 9 |
| Other | 73 | 74 |
| Not given | 243 | 205 |
